# Augmenting Deep Learning-Based PSMA PET/CT Metastasis Segmentation With a Population-Level Spatial Atlas

**DOI:** 10.64898/2026.08.26.26361439

**Authors:** Grant Chau, Bashirul Azam Biswas, Biratal Raj Wagle, Matthew E. Maeder, James B. Yu, Indrani Bhattacharya

**Affiliations:** Department of Computer Science Dartmouth College; Department of Biomedical Data Science, Geisel School of Medicine at Dartmouth, Hanover, NH 03755, USA; Radiology, Dartmouth Hitchcock Medical Center, Lebanon, NH 03766, USA; Radiation Oncology, Dartmouth Hitchcock Medical Center, Lebanon, NH 03766, USA

## Abstract

Automated lesion segmentation is increasingly central to PSMA PET/CT interpretation, supporting staging, treatment planning, and response assessment at a scale that outpaces available nuclear-medicine expertise. However, automated PSMA-PET/CT whole-body lesion segmentation models are trained on images alone, with no knowledge of where in the body prostate metastases actually tend to occur. Radiologists use clinical domain knowledge of metastatic spread, but its absence in machine learning models produces false positives in anatomically implausible locations and missed lesions in high-risk sites such as the liver. In this work, we explore whether population-level spatial knowledge of metastatic spread can be used to augment deep learning segmentation predictions, and how such a prior should be fused with a network’s output, without additional training. We build a data-driven metastasis atlas from 375 expert-annotated whole-body PSMA PET/CT scans and investigate its fusion with a trained segmentation network under a Bayesian framework, in which prediction probabilities from an nnU-Net-based lesion segmentation model serve as the likelihood and the data-driven atlas as the prior. Because metastases occupy only a small fraction of whole-body voxels, the atlas’s peak probability is too low, and standard power-scaled or naive Bayesian pooling references lack the tools to deal with this shortcoming. This causes these standard fusion strategies to fail and, in the naive Bayesian case, to sharply degrade performance. We instead derive a calibrated, background-referenced log-odds fusion, one of many possible approaches to combine a population atlas with a deep learning model’s predictions, distinct from classical multi-atlas label fusion in that it fuses a single population prior with a trained network’s softmax rather than combining several registered atlases. Furthermore, this approach is neutral outside atlas support by construction, reduces exactly to the baseline network when unweighted, and requires no retraining. This atlas fusion significantly improved mean Dice over the baseline nnU-Net on a disjoint internal test set (+0.011, Holm-adjusted *p* = 0.021) and on an independent external cohort (+0.0129, Holm-adjusted *p* = 3.8×10^*−*16^), with lesion sensitivity improving from 0.849 to 0.861 internally and Dice improving over baseline in every stratified anatomic region, including the rare, high-risk sites motivating this work, while naive Bayesian pooling degrades performance sharply and power-scaled pooling underperforms it throughout. Our findings suggest that population-level spatial priors can meaningfully augment deep learning predictions in whole-body oncologic segmentation, provided the fusion rule is calibrated to where the prior actually carries signal.

## 1 Introduction

Prostate cancer is the second leading cause of cancer death among American men, with metastasis responsible for most fatalities. Its incidence has risen since the mid-2000s [7], and an estimated 192,500 American men will be living with metastatic disease by 2030 [8]. Once it spreads, five-year survival falls from ∼ 100% in localized disease to ∼ 30% [1], with substantial associated morbidity [30, 39]. Prostate-specific membrane antigen positron emission tomography/computed tomography (PSMA PET/CT) and hybrid PET/magnetic resonance imaging (MRI) depict lymph-node and bone metastases more reliably than conventional cross-sectional imaging [10], and this detection role has grown to include selecting patients for PSMA-targeted radioligand therapy. For instance, the phase 3 VISION trial established that ^177^Lu-PSMA-617 prolongs survival in PSMA-positive metastatic castration-resistant disease [38], and the TheraP trial, whose protocol was designed to compare its activity and safety against chemotherapy [24], later demonstrated its efficacy relative to cabazitaxel [23], both using PSMA PET/CT to confirm eligibility.

Tumor burden computed directly from PSMA PET segmentations has itself become a clinically meaningful output to assess treatment response and as a prognostic metric. Furthermore, a bicentric analysis of 110 patients showed that total tumor volume, total lesion uptake, and their ratio, each computed from delineated lesions, predict overall survival after ^177^Lu-PSMA therapy [40]. Since total tumor burden computation depends on how well lesions are segmented, Kim et al. found in 78 patients that the choice of thresholding algorithm used to delineate lesions changes the resulting survival prediction [29]. Rios-Sanchez et al. used an automated total-tumor-volume computation pipeline for PSA50 response (at least 50% decline in baseline Prostate Specific Antigen PSA at any point after treatment initiation) prediction in ^177^Lu-PSMA-617 therapy, flagging segmentation errors within physiologically PSMA-avid organs as a specific limitation [34]. Thus, a burden estimate is only as trustworthy as the underlying segmentation. Seifert et al. later moved from a single whole-body volume to organ-specific volumes, quantified by a trained neural network across liver, lymph-node, bone, and other sites, and built a prognostic risk score from these region-level burdens in 1,348 patients [41], and Zang et al. showed that a fully automated, Artificial Intelligence (AI)-derived volumetric burden assessment predicts survival after ^177^Lu-PSMA-617 without any manual delineation [47].

A related line of work pairs radiomic features extracted from segmented lesions with clinical variables to predict treatment response rather than survival outright. Gutsche et al. trained a random forest model on radiomic features from segmented bone metastases to separate short-from long-term responders [19]. Moazemi et al. derived a radiomic signature from 83 patients’ pretherapeutic PET predictive of overall survival [33], and Roll et al. did the same from PET-MRI in 21 patients, predicting biochemical response [36]. Gong et al. combined multi-tracer imaging, including choline PET, with blood parameters such as neutrophil count and alkaline phosphatase to separate patients who fully benefit from radioligand therapy from those who do not [18], and Ghaderi et al. trained a multi-modal deep learning model directly on lesion-level imaging features from 99 patients to predict PSA progression-free survival [16].

These segmentation- and radiomics-derived predictors have in turn been formalized into nomograms and deployed clinical tools. Herrmann et al. built multivariable nomograms for overall survival, radiographic progression-free survival, and PSA response from the 551 patients treated in the VISION trial’s radioligand-therapy arm, with whole-body SUVmax among the imaging inputs [21]. Gafita et al. developed and externally validated similar nomograms on 196 development and 74 independent validation patients across six centers using earlier phase 2 data [12], and a version of this kind of tool is now deployed for clinical use by the UCLA nuclear medicine program [42]. However, PSMA-PET/CT interpretation suffers from high false positives, missed lesions, and inter-reader variability (*κ* = 0.41– 0.76) [9, 32], motivating automated segmentation whose anatomic fidelity is relevant for both detection and the down-stream burden estimates these clinical tools depend on.

Existing automated methods include nnU-Net [26] and PSMA-specific extensions using self-supervised representations, anatomy-aware pipelines, and cross-tracer adaptation [13, 14, 27, 28, 35], alongside transformer-based architectures such as Swin UNETR that have shown strong performance on related whole-body segmentation tasks [20].

None of these, however, encode where in the body prostate cancer actually spreads, costing them lesions in rare but high-risk sites like the liver and adding false positives (FPs) in anatomically implausible locations. Data-driven priors have improved detection elsewhere in prostate imaging [4, 31] and in segmentation more broadly, from probabilistic atlases [2] to learned CNN priors [6] and zonal information in modern networks [44].

To address this gap, we explore whether a population-derived, patient-specific spatial atlas of PSMA-avid metastases can be used as prior knowledge to augment deep learning segmentation, and if so, how it should be fused with a network’s predictions. This is one instance of the broader question of what population-level clinical knowledge can be productively fused with deep learning predictions. Specifically, our contributions are:

- We construct a population-derived, whole-body spatial atlas of PSMA-avid metastases from 375 expert-annotated PSMA PET/CT scans, and use it as a spatial prior for deep learning-based lesion segmentation.
- We show that the standard way of fusing a prior with a network’s prediction, naive Bayesian pooling and its refinement, power-scaled pooling, both treat 0.5 as the point where the atlas has “no opinion”. Because our atlas’s probabilities never get close to 0.5 anywhere in the body, that assumption doesn’t hold for a sparse atlas like ours, and naive Bayesian fusion in particular sharply degrades performance as a result.
- We derive a calibrated, background-referenced log-odds fusion rule (Eq. 2) that instead treats the atlas’s own background level as its “no opinion” point, requires no retraining, and reduces exactly to the baseline network when unweighted.
- We demonstrate that this atlas fusion significantly and reproducibly improves Dice over a strong nnU-Net baseline on both a held-out in-distribution test set and an independent out-of-distribution cohort, surviving multiplicity correction.
- We evaluate performance at the voxel, lesion, and anatomic-region level, and show that atlas fusion improves Dice in every anatomic region evaluated, including the rare, high-risk sites (liver, lung, high-risk bone) that motivate this work.

## 2 Materials and Methods

Figure 1 summarizes our approach. The pipeline has two phases. Atlas construction (Fig. 1a-b) happens once, offline, using only the training partition: each training subject’s CT is registered to a common reference frame, and the resulting warped lesion masks are averaged voxel-wise into a single population atlas of where PSMA-avid metastases tend to occur in the body. At inference (Fig. 1c-d), that fixed atlas is warped into each new patient’s own anatomy and combined, voxel by voxel, with the trained segmentation network’s softmax prediction through the calibrated log-odds pooling rule derived in Sec. 2.4. Because the atlas is built once and reused unchanged across patients, no additional training or per-patient atlas construction is required at test time.

**Figure 1.**
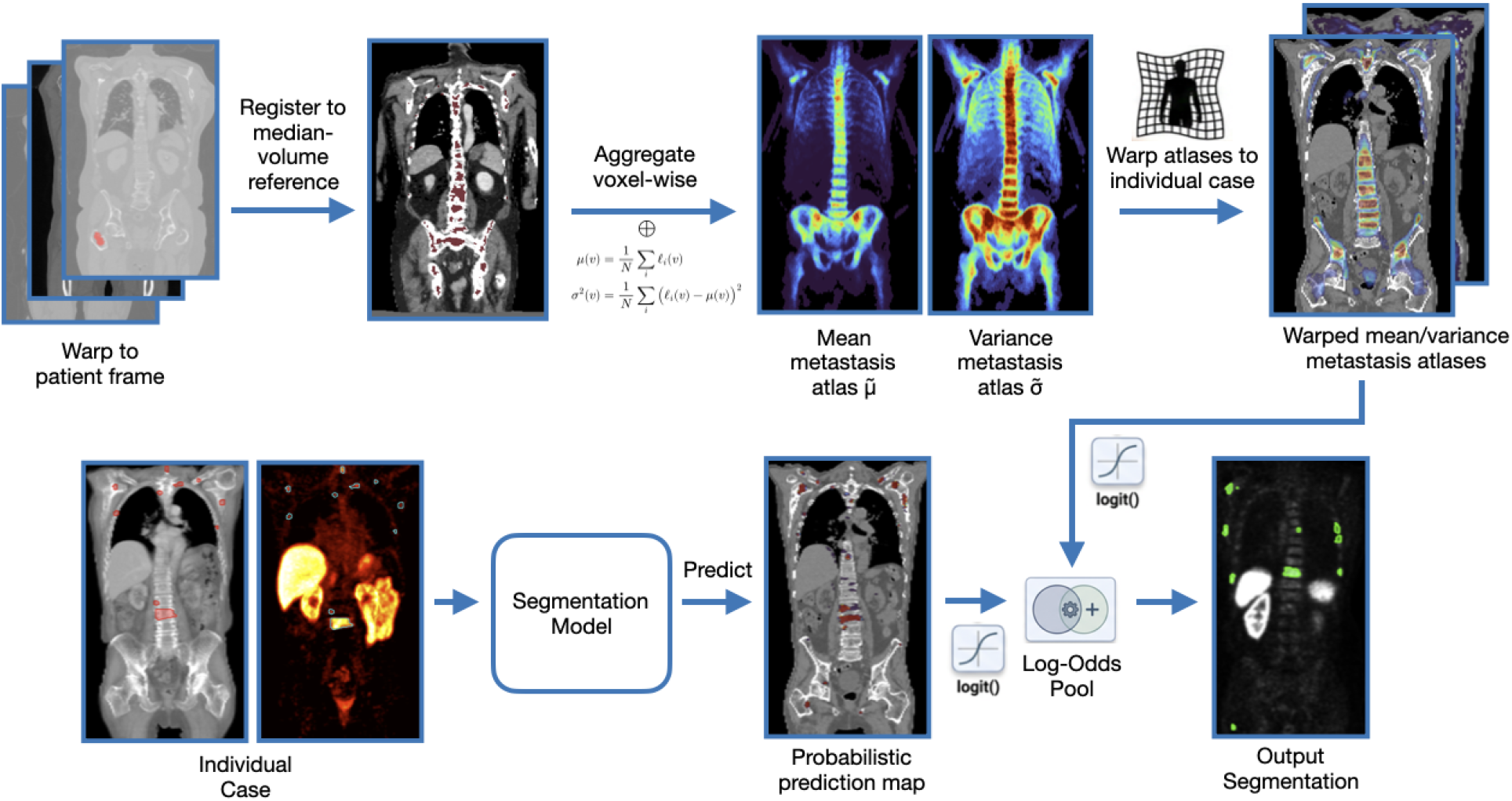
Method overview. A population atlas is built once from the 375-subject training partition by registering each subject’s CT to a median-volume reference (SyN) and warping the binary lesion masks into that space **(a)**, then aggregating them voxel-wise into the mean atlas *µ*(*v*) **(b)**. At inference the atlas is warped into the patient’s frame **(c)** and fused with the nnU-Net softmax through the calibrated log-odds pool of Eq. 2 **(d)**.

### 2.1 Data and baseline training

We used the PSMA subset of the publicly available AutoPET III dataset [27] and, as an independent out-of-distribution cohort, the PSMA subset of the Deep-PSMA dataset. Table 1 summarizes these two cohorts, reproduced from the corresponding PSMA columns of the pan-cancer, multi-tracer dataset summary. AutoPET-III PSMA cases were partitioned once at the subject level, prior to any modeling, into disjoint training (379 scans, 343 disease-positive), validation (96, 84) and test (122, 110) sets. The atlas and baseline network were estimated only from the training partition, so no validation or test voxel informed the prior or the network. All results are on disease-positive cases. The baseline is a 3D full-resolution nnU-Net [26] trained from scratch on the training partition with the framework’s default self-configuring hyperparameters (SGD, Nesterov momentum 0.99, Dice + cross-entropy loss, initial LR 0.01 with polynomial decay over 500 epochs, and standard augmentation). Softmax probabilities were retained at inference for log-odds combination.

**Table 1:** Dataset summary for the two PSMA cohorts used in this study (reproduced from the PSMA columns of the pan-cancer, multi-tracer dataset summary in [45]). Pr=Prostate, Neg=Negative, *dz* = distance, *µ*_*PET*_ = Mean PET Intensity.

| Properties | AutoPET-III (PSMA) | Deep-PSMA (PSMA) |
| --- | --- | --- |
| # Patients | 597 | 100 |
| Sex (M/F/NS) | 597/0 | 100/0 |
| Disease dist. (Pr/Neg) | 537/60 | 100/0 |
| #lesions (per case) | $35.04 \pm 51.70$ | $68.41 \pm 58.35$ |
| $\mu_{PET}$ (per lesion) | $7.48 \pm 7.12$ | $5.64 \pm 3.33$ |
| # slices (per vol) | 135–963 | 195–1261 |
| In-plane res. (mm) | 2.73–4.07 | 1.59–5.47 |
| $dz$ bet. slices (mm) | 2.00–5.00 | 1.5–5.00 |
| train/val/test | 375/96/122 | 0/0/100 |

**Table 2:** Ablation configurations compared throughout this study. 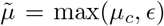 is the atlas prior with numerical floor *ϵ* = 10^*−*6^; operating points (*α, λ*) were selected on the validation cohort and then frozen for all reported test-set results.

| Method | Formula | Parameters | Description |
| --- | --- | --- | --- |
| Baseline | $\hat{P} = p$ | — | Raw nnU-Net softmax prediction; no atlas information is used. |
| Naive Bayesian fusion | Eq. 1, $\alpha = 1$ | $\alpha = 1$ | Multiplies the network probability by the atlas prior directly; implicitly treats $\tilde{\mu} = 0.5$ as the atlas’s “no opinion” point. |
| Power-scaled pooling | Eq. 1 | $\alpha = 0.055$ | Shrinks the prior toward that same 0.5 point with exponent $\alpha$ before pooling. |
| **Atlas fusion (ours)** | Eq. 2 | $\alpha = 0.25, \lambda = 0.8$ | References the prior to its own background floor $\epsilon$ rather than 0.5, with separately tunable shape ($\alpha$) and strength ($\lambda$). |

### 2.2 Probabilistic model

We treat segmentation as voxelwise inference of a label *Y* from evidence *X* given disease status *C*, under the factorization 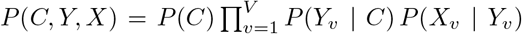, where *v* = 1, …, *V* indexes voxels of the whole-body PET/CT volume, labels are conditionally independent given *C*, and evidence depends only on the local label. These assumptions are approximations, as contiguous lesions violate spatial independence. However, they yield a closed-form posterior matching how the network and atlas are estimated. We identify the conditional prior *P* (*Y*_*v*_ = 1 | *C* = 1) with the warped atlas value *µ*_*c*_(*v*) and the likelihood ratio *P* (*X*_*v*_ | *Y*_*v*_ = 1)*/P* (*X*_*v*_ | *Y*_*v*_ = 0) with the network odds *p/*(1 *− p*), where *p*(*v*) is the softmax foreground probability.

### 2.3 Atlas construction

We first select the cohort subject with median body volume as the **reference** registration target, minimizing average deformation. Each subject’s CT was **registered** to the reference by affine initialization then symmetric diffeomorphic (SyN) deformation with a cross-correlation metric [3] (ANTs SyNCC), giving transform *T*_*i*_. Applying *T*_*i*_ to the binary lesion mask (nearest-neighbor) yields *l*_*i*_ in reference space. The **voxelwise mean** 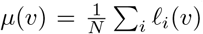 over *N* =375 maps forms the marginal atlas. The marginal *µ*(*v*) averages over disease status; since lesions occur only in positive cases, *P* (*Y*_*v*_=1) = *P* (*Y*_*v*_=1|*C*=1) *P* (*C*=1) with *P* (*C*=1) *≈ N*_+_*/N*, so dividing by the positive fraction recovers the conditional prior: *µ*_*c*_(*v*) = min(1, *µ*(*v*)· *N/N*_+_), *N/N*_+_ = 375*/*332 = 1.13, giving *µ*_*c*,max_ ≈ 0.18. Inference 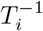 maps *µ*_*c*_ onto the patient grid by linear interpolation (the nearest neighbor would inject plateaus). Figure 1b shows the resulting mean atlas.

### 2.4 Power-scaled pool and log-odds fusion

We combine two independent pieces of evidence about whether a voxel contains a lesion: the network’s prediction *p* and the atlas prior *µ*_*c*_. A standard way to combine two probabilistic opinions is to add their log-odds 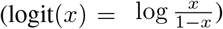, a technique known as an opinion pool [5, 22]: a source favoring foreground raises the combined score, one favoring background lowers it. What remains is how to weight and calibrate the atlas’s contribution.

The simplest pool multiplies *p* and *µ*_*c*_ (naive Bayesian fusion), which implicitly sets each source’s neutral, “no opinion” point at 0.5, since logit(0.5) = 0. Our atlas never reaches that threshold anywhere in the body: even at its most metastasis-prone voxel *µ*_*c*,max_≈ 0.18 (Sec. 2). Multiplying by a value always below 0.18 therefore pulls every voxel toward background, including lesions the network already detects correctly, and indeed naive Bayesian fusion deteriorates Dice from 0.517 to 0.465 and lesion sensitivity from 0.849 to 0.782 (Table 3). The standard fix shrinks the prior with an exponent *α ∈* [0, 1] (a “power-scaled” or log-linear pool) [11, 15]:

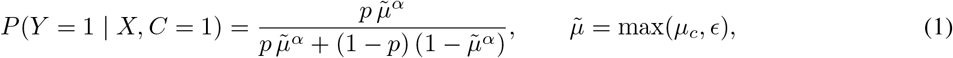

with numerical floor *ϵ* = 10^*−*6^. The 0.5 neutral point persists here, since the prior term reinforces foreground only when 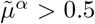. Our fusion rule instead references the prior to its own floor *ϵ*, the level voxels take where there is no sign of disease spread, so the atlas pushes toward foreground only where a voxel is genuinely more metastasis-prone than typical background:

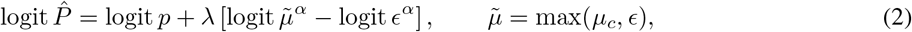

Here *α* shapes how the prior’s contribution grows as *µ*_*c*_ rises above background, and *λ* sets how strongly that comparison is allowed to shift the network’s prediction. This gives two useful properties directly from the formula: at any voxel with no atlas support (*µ*_*c*_ ≤ *ϵ*), the bracket is zero, so the prediction is left exactly as the network gave it 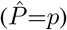; and setting *λ*=0 recovers the network’s prediction everywhere, regardless of *α*. The shape of the prior’s influence (*α*) and its overall strength (*λ*) can therefore be tuned independently, and the atlas can never push a background voxel toward the foreground on its own.

### 2.5 Ablation Studies

To isolate the effect of each modeling choice, whether an atlas is used at all, how heavily it is weighted, and where its pooling rule anchors the atlas’s “no opinion” point, we compare four configurations throughout Tables 3–5. Table 2 summarizes them.

**Table 3:** Voxel- and lesion-level performance on the AutoPET PSMA in-distribution test set (*N* =110 disease-positive, 3,231 ground-truth lesions). FN/FP Vol are per-case means in cubic cm.

| Method | Dice | FN Vol Mean | FP Vol Mean | Lesion Sensitivity | PPV |
| --- | --- | --- | --- | --- | --- |
| Baseline | 0.517 | 9.530 | 15.234 | 0.849 | 0.778 |
| Naive Bayesian Fusion ($\alpha = 1$) | 0.465 | 13.626 | **8.025** | 0.782 | **0.861** |
| Power-scaled ($\alpha = 0.055$) | 0.526 | 9.167 | 16.837 | 0.859 | 0.764 |
| **Atlas fusion** ($\alpha = 0.25, \lambda = 0.8$) | **0.528** | **9.126** | 17.354 | **0.861** | 0.757 |

**Table 4:** Performance stratified by anatomic site on the AutoPET PSMA in-distribution test set (liver, lung, bone, and high-risk bone [HR-bone] subregions). Dice/FN/FP are row-level means over TotalSegmentator label instances with overlap *>*0.4 cm^3^ (liver: *>*2.0 cm^3^, given confounding physiological uptake). FN/FP Vol in cm^3^.

| Method | Liver |  |  | Lung |  |  | Bone |  |  | HR-bone |  |  |
| --- | --- | --- | --- | --- | --- | --- | --- | --- | --- | --- | --- | --- |
|  | Dice | FN Vol | FP Vol | Dice | FN Vol | FP Vol | Dice | FN Vol | FP Vol | Dice | FN Vol | FP Vol |
| Baseline | 0.206 | 1.882 | 31.176 | 0.350 | 0.907 | 0.218 | 0.614 | 0.338 | 0.723 | 0.626 | 0.322 | 1.205 |
| Naive Bayesian | 0.075 | 15.662 | **11.958** | 0.143 | 1.530 | **0.032** | 0.535 | 0.478 | **0.483** | 0.566 | 0.498 | **0.842** |
| Power-scaled | 0.211 | 1.800 | 33.430 | 0.375 | **0.788** | 0.243 | 0.621 | 0.309 | 0.811 | **0.630** | 0.304 | 1.340 |
| **Atlas fusion** | **0.212** | **1.772** | 34.266 | **0.380** | 0.792 | 0.251 | **0.622** | **0.307** | 0.839 | **0.630** | **0.300** | 1.382 |

**Table 5:** DeepPSMA out-of-distribution test-cohort metrics (N =100), parameters frozen from AutoPET validation (no recalibration on DeepPSMA). Dice_µ±σ_ is the paired mean ±SD, consistent with ΔDice, which is also mean-based. ΔDice is the paired mean vs. baseline with bootstrap 95% CI. p is Holm-adjusted Wilcoxonover the three comparisons.

| Method | $\text{Dice}_{\mu \pm \sigma}$ | $\text{Dice}_{\text{med}}$ | $\Delta\text{Dice}_{\mu}$ (95% CI) | $p$ (Holm) |
| --- | --- | --- | --- | --- |
| Baseline | $0.600 \pm 0.157$ | 0.648 | — | — |
| Naive Bayes ($\alpha = 1$) | $0.464 \pm 0.147$ | 0.492 | $-0.136 [-0.160, -0.113]$ | $9.56 \times 10^{-15}$ |
| Power-scaled ($\alpha = 0.055$) | $0.607 \pm 0.157$ | 0.650 | $+0.007 [0.006, 0.008]$ | $9.56 \times 10^{-15}$ |
| **Atlas fusion** ($\alpha = 0.25, \lambda = 0.8$) | **$0.613 \pm 0.158$** | **0.655** | **$+0.013 [0.011, 0.015]$** | **$3.82 \times 10^{-16}$** |

### 2.6 Evaluation Methods

#### 2.6.1 Voxel-level Evaluation

Voxel-level evaluation captures overall volumetric overlap between predicted and reference lesion masks, independent of how many discrete lesions are involved. We report Dice (median, mean ±SD), computed only on disease-positive cases, together with false positive volume (FPVol) and false negative volume (FNVol) in cubic centimeters. Lesion masks were converted to discrete connected components using 18-connectivity. Furthermore, predictions with zero overlap against every ground-truth lesion were scored a Dice of 0 rather than excluded, so that missed cases are penalized rather than dropped from the average. Paired differences between the baseline and each fusion strategy were assessed using bootstrap confidence intervals and the Wilcoxon signed-rank test with Holm correction [25], alongside the matched-pairs rank-biserial effect size *r*_*rb*_ and Two One-Sided Tests (TOST) against a 0.01 Dice non-inferiority margin. The primary endpoint is the paired mean Dice difference against the prior-off baseline on the held-out test set. Holm correction is applied once across the three fusion strategies compared. All reported *p*-values are Holm-adjusted and compared against *α* = 0.05.

#### 2.6.2 Lesion-level Evaluation

Voxel-level Dice can mask clinically important behavior: a method that enlarges an already-detected lesion and a method that finds an entirely new one can produce the same Dice gain, but only the latter changes clinical management. Lesion-level evaluation instead asks how many discrete lesions were found, independent of how tightly their boundaries were drawn. We converted both ground-truth and predicted voxel masks into discrete lesions via 3D connected-component analysis; a ground-truth lesion was scored as detected if any predicted voxel overlapped it, and predicted components with no corresponding ground-truth lesion were scored as false positives. Sensitivity and positive predictive value (PPV) were computed micro-averaged, i.e., by pooling all lesions across all test-set patients before computing the ratio, rather than averaging per-patient rates, so that patients with many lesions are not down-weighted relative to patients with few. Lesion count is directly relevant to treatment planning: in oligometastatic prostate cancer, where disease is limited to a small number of metastatic sites, aggressive metastasis-directed therapy has been shown to improve outcomes in this heterogeneous, poor-prognosis population [45], making an accurate lesion count, not just an accurate lesion boundary, clinically consequential.

#### 2.6.3 Anatomic Region-level Evaluation

Not all missed or spurious lesions carry equal clinical weight: a missed liver metastasis or a missed high-risk bone lesion can change a patient’s management in ways that a missed low-risk bone lesion would not. We therefore additionally stratify performance by anatomic region. CT-based anatomic structures (liver, lung, and bone) were delineated using the pretrained TotalSegmentator model [46], and Dice, FNVol, and FPVol were recomputed restricted to voxels within each structure. Following the high-risk bone metastasis criteria from a multicenter randomized trial of prophylactic radiotherapy [17], also used in our companion study [45], we additionally investigate how well our method detects **high-risk bone** metastases. High-risk bone metastases are those meeting at least one of: bulky (≥2 cm) disease, involvement of the hip, shoulder, or sacroiliac joint, disease in long bones occupying one-third to two-thirds of cortical thickness, or disease in the junctional spine (C7–T1, T12–L1, L5–S1) or with posterior element involvement [17]. These criteria mark bone lesions where symptomatic progression or skeletal-related events are more likely, and where segmentation errors are therefore more consequential for treatment planning than in bone disease generally.

### 2.7 Results

#### Parameter selection

Both selection stages used the validation cohort only. Sweeping *α* ∈ [0.005, 1.0] with *λ* fixed, mean voxel Dice flattens at 0.531 over *α*∈ [0.08, 0.15] (baseline 0.529), an interior optimum that rules out both *α* →0 (prior dominates) and *α*→ 1 (naive Bayesian fusion), consistent with the sign reversal of Sec. 2.4. A joint grid search over (*α, λ*) gave the optimum *α* = 0.25, *λ* = 0.8 (*λ* = 0 recovers the baseline exactly). This operating point was then frozen and applied unchanged to the held-out test set and the out-of-distribution DeepPSMA cohort, so any gain there cannot be attributed to overfitting the operating point.

#### Voxel-level results

On the in-distribution test set (Table 3), atlas fusion’s paired mean Dice gain (+0.011, Holm *p*=0.021) exceeded power-scaled’s (+0.009, *p*=0.036). The same ordering holds on the out-of-distribution DeepPSMA cohort (Table 5): atlas fusion gives the largest Dice gain of any strategy (+0.0129, Holm *p* = 3.8× 10^*−*16^), while naive Bayesian fusion again degrades sharply (−0.1362). Unlike power-scaled, whose gain there is statistically equivalent to a trivial (*<* 0.01 Dice) shift by TOST (*p* = 1 × 10^*−*6^), atlas fusion’s is not (*p* = 0.9996), consistent with its larger effect size (*r*_*rb*_ = 0.95 vs. 0.89).

#### Lesion-level results

The Dice gain reflects an additive pool enlarging existing correctly-located components rather than creating new ones. Atlas fusion increases lesion sensitivity from 0.849 to 0.861 and detects 0.35 more lesions per case, at a small precision cost (PPV 0.778→ 0.757) common to all priors tested (Table 3). Figure 2 shows this in three lesions the baseline left below threshold, two recovered outright and one tightened at the boundary. Naive Bayesian fusion instead collapses both Dice and detected-lesion count despite a lower FP volume, so its lower false-positive rate comes from suppressing true positives rather than correcting false ones.

**Figure 2.**
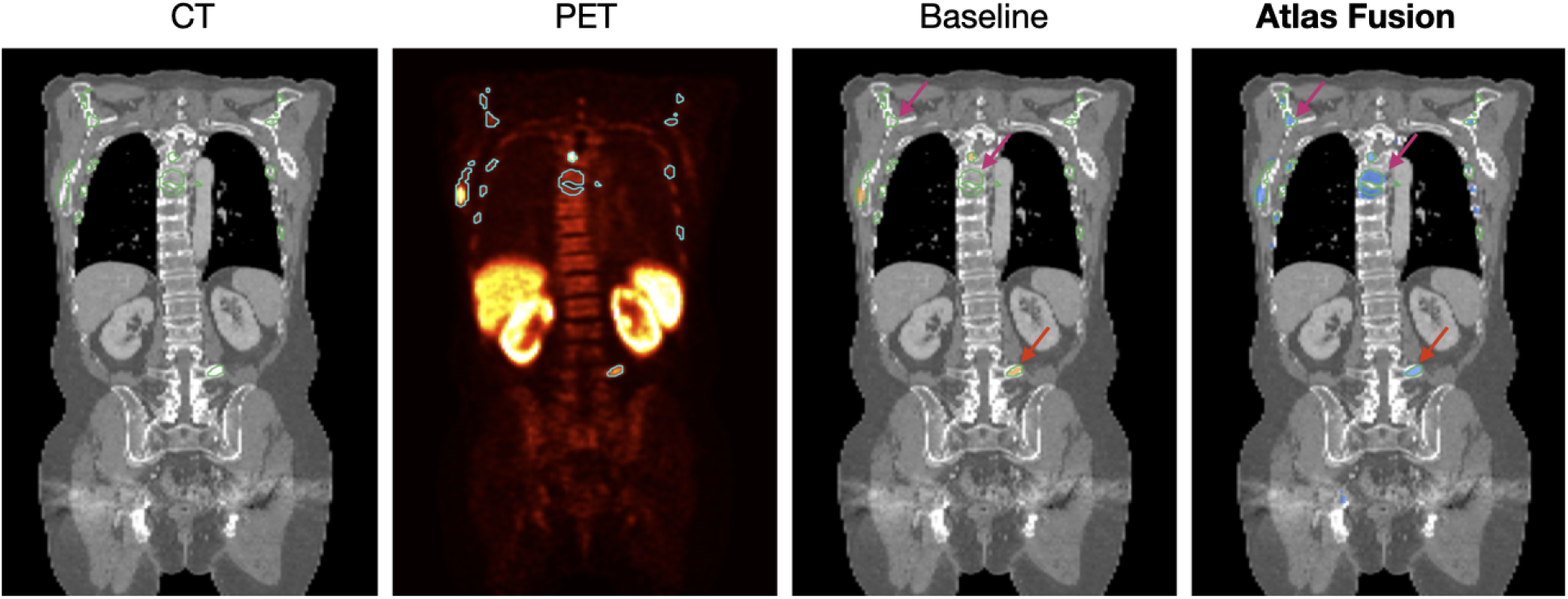
Qualitative lesion recovery. The first two columns show the coronal CT and PET scans with outlines of cancer labels, while the third and fourth columns show the baseline and atlas fusion predictions, respectively. Baseline nn-UNet misses a 0.52 cm^3^ vertebral metastasis and a 0.92 cm^3^ right scapular metastasis (Magenta arrows), that atlas fusion detects. Baseline nn-UNet partially detects a smaller verterbal lesion (red arrow), which atlas fusion delineates with a tighter boundary. PET (second column) is visualized to confirm tracer uptake.

#### Anatomic region-level results

Stratified by site (Table 4), this same pattern holds most sharply in the rare, high-risk sites motivating this work: naive Bayesian fusion collapses Dice in liver and lung, while atlas fusion improves Dice over baseline in every region evaluated (liver, lung, bone, and high-risk bone).

#### Qualitative results

Figure 2 illustrates these voxel- and lesion-level gains in three representative cases from the held-out test set. Baseline nnU-Net misses two lesions entirely, a 0.52 cm^3^ vertebral metastasis and a 0.92 cm^3^ right scapular metastasis, both recovered by atlas fusion; the corresponding PET images confirm tracer uptake at both sites, ruling these out as spurious atlas-driven additions rather than genuine detections. A third, smaller vertebral lesion is only partially detected by the baseline and is delineated with a tighter boundary under atlas fusion, showing that the fusion rule both recovers lesions the baseline missed outright and sharpens boundaries on lesions it already found. These cases are consistent with the aggregate lesion-level statistics above: gains concentrate in lesions the baseline placed just below its detection threshold, at locations the atlas marks as spatially plausible.

## 3 Discussion and Conclusion

This work set out to explore whether population-level knowledge of where prostate cancer metastases occur can augment deep learning segmentation of PSMA PET/CT. This work represents one way of injecting clinical prior knowledge into an otherwise image-only model and how such a prior should be fused with a network’s predictions.

We find that it can, and the calibrated, background-referenced log-odds pool this yields fuses a population-derived whole-body metastasis atlas with any pretrained network’s softmax at inference, without retraining. This atlas fusion improves Dice reproducibly (+0.002, +0.011, +0.0129 from validation to in-distribution to out-of-distribution test) and in every stratified anatomic region, most notably the rare, high-risk sites (liver, lung, high-risk bone) that motivated this work. Examining the tails shows why: gains recover lesions the baseline left below threshold (+0.35/case), while losses arise when the same additive boost reaches false positives in high-prior regions, where physiological uptake in the axial skeleton is indistinguishable from true lesions. Because *λ* and *α* set this trade explicitly, the operating point can be re-tuned toward sensitivity or precision as the use case demands. Requiring no retraining and no architecture-specific integration, this fusion rule can be dropped onto any pretrained model at negligible compute cost. Notably, a preliminary attempt to instead inject the atlas as a retrained input channel underperformed this inference-only formulation.

We evaluated a single baseline architecture and one registration pipeline, and our in-distribution test cohort shares an acquisition source with training. The DeepPSMA cohort addresses the latter directly. Extending this approach to additional architectures, such as transformer-based segmentation networks like Swin UNETR [20], to other registration methods, and to further scanner sources remains future work. Longitudinal tracking of individual lesions across serial scans is a related extension the atlas could support directly, since a patient-specific atlas warp already places each scan in a common reference frame. Automated multimodal lesion-tracking methodology [37] could build on this directly.

PSMA-PET/CT is transforming advanced prostate cancer care through more accurate staging, guiding treatment decisions, disease progression tracking, and outcome prediction. Missing a liver metastasis or a high-risk bone lesion is not a benign segmentation error, as liver involvement carries a substantially worse prognosis than nodal or non-high-risk bone spread alone, and an undetected high-risk bone lesion forgoes the window for metastasis-directed radiotherapy that can prevent skeletal-related events such as pathologic fracture or cord compression [17]. These are also exactly the sites where automated segmentation struggles most, consistent with the missed lesions and inter-reader disagreement reported more broadly for PSMA PET/CT interpretation [9, 32]: baseline nnU-Net’s Dice in our own results falls to 0.206 in liver, far below its whole-body Dice of 0.517 (Table 4). Moreover, since tumor burden derived from PSMA-PET segmentation increasingly feeds prognostic and treatment-response models for PSMA-targeted radioligand therapy, whose use is expanding earlier in the disease course [43], we see anatomically informed segmentation of this kind as relevant beyond voxel-level Dice. The organ-specific risk score of Seifert et al. is a particularly close analog to our own anatomic-region stratification, since it derives separate prognostic value from lymph-node and bone tumor volume [41], which closely represent the regions where atlas fusion changed Dice in Table 4. Atlas-referencing the segmentation step could potentially change the total-tumor-volume estimates feeding pipelines like that of Zang et al. [47] or the earlier bicentric analysis of Seifert et al. [40]. Furthermore, whether applying atlas on the imaging inputs anchoring the VISION-trial nomograms of Herrmann et al. [21] and the phase 2 nomograms of Gafita et al. [12] is an open question for future work. The resulting framework is a tunable foundation for operating-point selection, richer non-marginal priors, and, more broadly, exploring what other forms of population-level clinical knowledge can be productively fused with deep learning predictions in settings like this one.

## Data Availability

PSMA-PET-CT-Lesions / AutoPET PSMA (https://doi.org/10.7937/R7EP-3X37)
Deep-PSMA dataset link (https://zenodo.org/records/15281784)

https://doi.org/10.7937/R7EP-3X37

## Acknowledgments

Research reported in this publication was supported by an Institutional Development Award (IDeA) from the National Institute of General Medical Sciences of the National Institutes of Health under grant number 1P30GM149408. We also gratefully acknowledge support from the Munck-Pfefferkorn Fund, the Center for Molecular Epidemiology Pilot, and the American Cancer Society IRG at the Geisel School of Medicine. We furthermore thank the Departments of Computer Science and Biomedical Data Science at Dartmouth College, as well as the Department of Radiology at Dartmouth Hitchcock Medical Center. All of this support was crucial in making this work possible. Help from Claude Sonnet 5 was taken for correcting grammatical errors.

